# HIV risk perception and self-reported risk behaviors among men who have sex with men using social media in Beijing, China and its implications on HIV testing

**DOI:** 10.1101/2022.11.23.22282660

**Authors:** Yafang Zhao, Daniel J Bromberg, Kaveh Khoshnood, Alec Dai, Zhen Li, Yu Sheng

## Abstract

Inaccurate HIV risk perception is common among men who have sex with men (MSM). Accurate self-perceived risk and HIV testing are closely related and are essential in promoting HIV treatment cascade succeed. This cross-sectional study aims to 1) explore HIV risk perception and its associated sexual behaviors, 2) identify associated factors with HIV testing among Chinese MSM. Participants were recruited to an anonymous online survey in 2019 via an MSM social application. The questionnaire included sexual behaviors, HIV testing history, and the use of HIV prevention services. Logistic regression was used to explore the factors associated with higher risk perceptions and HIV testing in the past 12 months. Total 431 MSM were recruited, 73.3% had been tested for HIV in the past year and 47.80% of MSM self-reported in the high-risk group. MSM who perceived a higher risk (AOR=3.42, 95% CI:2.06-5.68), had multiple sexual partners (AOR=2.60, 95% CI:1.40-4.83), knew sex partner’s HIV status (AOR=7.96, 95% CI:4.33-14.65), and have STI diagnosis (AOR=2.76, 95% CI:1.10-6.95), were more likely to have been tested. Risk perception and sexual behavior were associated with HIV testing. Culturally adapted, theory-driven education programs are expected to utilize in improving MSM’s risk perception accuracy and HIV testing.

## Introduction

In Asia, the number of men who have sex with men (MSM) with the human immunodeficiency virus/acquired immunodeficiency syndrome (HIV/AIDS) has dramatically increased over the past decade[1]. The prevalence of HIV via male-to-male sexual transmission in China has rapidly risen from 6.1% in 2011 to 9.2% in 2017[2, 3]. According to three separate cohort studies among MSM in Beijing, HIV incidence rates increased from 5.9 per 100 person-years in 2009[4] to 8.1 per 100 person-years in 2010[5]. It is necessary to identify MSM who are HIV positive to reduce HIV infections [6]. HIV testing is one cost-effective strategy[7] that increases the proportion of known HIV diagnoses, thus prompting the start of the treatment cascade. The Chinese government has launched a treatment as prevention (TasP) strategy[8] which recommends increasing HIV testing for MSM who have multiple sexual partners, anonymous sexual partners, or other behaviors that contributes to their risk of HIV infection.

HIV testing coverage is still insufficient among MSM globally [9]. A large pan-European survey among MSM named the European MSM Internet Survey (EMIS) 2010[10], and a follow-up survey among Germans in 2013[11], indicated that about a third of MSM have never been tested for HIV, more than a third of the participants who had never been diagnosed with HIV got HIV testing in the past 12 months, and less than a third were tested even longer ago. Testing is infrequent among MSM who have high risks of infection[12, 13]. According to one systematic review and meta-analysis focusing on HIV testing among Chinese MSM[14], fewer than 50% of Chinese MSM reported having ever been tested, and only 38% of Chinese MSM reported having been tested in the past 12 months.

The self-perceived risk of getting HIV has been shown as a major factor of whether MSM get HIV testing or not [15, 16]. Previous studies reported some risk behaviors related to HIV risk perception among MSM, such as the number of anal sex partners, condom use during sex, knowledge of the partner’s HIV status, and other sexual risk behaviors. These factors therefore can shape testing behavior[17]. Risk perception awareness among the MSM community is insufficient[16]. Over six months, Chinese MSM have, on average, 6.17 (3.45-9.96) sexual partners[18], and approximately 53% and 75% of MSM have had unprotected anal intercourse (UAI) according to two systematic meta-analyses conducted in China[19, 20]. Lack of sexual knowledge, high frequency of condomless anal intercourse, having sex in exchange for money/drugs, and other risky behaviors are prevalent among MSM and significantly increase their likelihood of getting HIV infections[21, 22]. Congruency between self-perception of HIV risk and reported risk-taking behaviors is especially important when thinking about increasing HIV testing[23, 24]. Stephenson et al.’s study[24] indicated a significant proportion of MSM rated their self-perceived HIV risk as low despite actually having high-risk behaviors. Therefore, accurate risk awareness is especially important in effectively promoting HIV testing and preventing HIV transmission among MSM.

A better understanding of risk perception and sexual risk behaviors associated with HIV testing behavior among MSM who are at an increased risk for HIV may help to devise improved and more effective strategies to promote HIV testing. Our study attempts to address some limitations of previous studies by 1) evaluating HIV risk perception and correlating risk behaviors among Chinese MSM; 2) analyzing the associations between HIV risk perceptions, self-reported risk behaviors, and HIV testing to identify the most relevant factors for improving HIV testing behavior among MSM in China.

## Materials and Methods

### Study Design

This cross-sectional study was conducted utilizing an anonymous voluntary online survey via a social application named BLUED.

### Ethics

This study was reviewed and monitored by the Institutional Review Boards of Peking Union Medical College. All participants completed the written informed consent.

### Participants

Users of BLUED who disclosed their sexuality as either gay or bisexual were the targeted population and categorized as MSM of this study. The participants were (a) 18 years and above, (b) self-identified as cisgender men, (c) had unprotected sex with at least one male in the past year, and (d) provided consent to participate in the survey. MSM who were a) HIV positive, b) diagnosed with severe mental health disorders (e.g., major depression disorder or anxiety disorder according to the DSM-5 system, such as dysthymia and double depression) were excluded.

It estimated that of all MSM who reported unprotected sex with male in the past 12 months in Changsha, only 62.3% had undergone testing for HIV [25]. We set the study power of at least 80%, the significant level at 5%, *d*=0.06, and considered a 30% drop off rate, we calculated a minimum sample size of 358 participants.

### Measures

The risk behaviors section of this online survey was adapted from the American Men’s Internet Survey (AMIS) which is a self-reported, annual Internet-based behavioral survey of MSM living in the United States[26]. Core questions include demographic characteristics, sexual behaviors, HIV testing history, and the use of HIV prevention services, etc. AMIS has been frequently utilized in many previous online surveys[27, 28]. More details of the questionnaire are presented in the **Supplementary document**.

#### (1) Sociodemographic Characteristics

Participants were asked to self-report demographic characteristics such as age, education level, age of first sexual intercourse, and sexual orientation.

#### (2) HIV Testing Behaviors

MSM who enrolled in the survey were asked about the history of HIV testing in their lifetime and in the past 12 months.

#### (3) HIV Risk Perception

Participants answered three short questions which have been reported in previous study[29]. All items were summed and recorded into a total score, ranging from 0-12. We categorized participants into two levels based on the summed score of the three questions (“low risk” referred to a score of ≤6, “high risk” referred to a score higher than 6). Internal consistency of the 3 items assessment was high (Cronbach’s α = .84).

#### (4) Risk Behaviors

Four aspects adapted from AMIS as follow: (1) sexual behavior with a male sex partner(s), (2) substance use, (3) sexually transmitted infections (STI) diagnoses, and (4) assessment of prevention activities.

### Procedures

We recruited participants from 10 July of 2019 to 26 July of 2019. The research team and a trained MSM peer were responsible for developing an online survey. Three questions for screening eligibility were provided before the consent document. Only people who met the criteria were able to review the consent form. After clicking to consent, they can start answering the main questionnaire. The URL of the questionnaire was sent by private message on BLUED to users whose ZIP address was in Beijing. Participants used their smartphones to take part in the survey and a unique IP address control was used to avoid duplicates or false entries on the link. Data was collected through a web platform (www.sojump.com).

### Statistical Analysis

We performed statistical analyses using SAS (Version 9.4; Cary, NC, USA.)[30]. Descriptive statistics for socio-demographics, HIV testing behavior, HIV risk perceptions, and risk behaviors were used to explore the distribution, central tendency, variance, proportions of categorical variables, and mean standard deviation for continuous variables.

Bivariate logistic regression was used to select the potential factors of higher risk perception (setting “low risk” as the reference group) from self-reported risk behaviors. For the second study aim (determining associated factors of HIV testing), bivariate logistic regression was conducted to identify sociodemographic factors, self-reported risk factors, and HIV risk perceptions associated with HIV testing in the past 12 months (“Yes” =1, “No” =0). Then, for multivariate analyses, factors from the bivariate analysis with a significant *p*-value were entered with a backward selection approach to create the final model. A *p*-value of < 0.05 with 95% confidence interval was considered as significant.

## Results

### Demographic Characteristics and HIV Testing Behaviors

A total of 1166 potential participants were recruited to participate in the study. 67 were excluded as they did not provide consent. 640 were further excluded due to self-reporting that they did not have unprotected sex with males in the past 12 months. Seven were excluded due to missing information on risk behaviors. Therefore, a total of 431 MSM were included in our final survey and analysis.

**Error! Reference source not found**.presents the results of socio-demographic and HIV testing behavior. Their average age was 31.1 years (SD=8.1), the average age of their first sexual intercourse was 20.4 years (SD=4.8). 78.2% (337) participants were single, 79.3% (340) identified their sexual orientation as gay. 73.3% of MSM had an HIV test in the past year.

### HIV Risk Perception, Self-Reported Risk Behaviors, and HIV Testing

Results of self-reported risk behaviors in each perceived risk subgroups were shown in **Error! Reference source not found**.. 47.80% (206/452) of MSM self-reported to be in the high-risk group. There was a significant difference (*p*< 0.05) in the number of the sexual partner(s) between low-risk and high-risk subgroups. Most MSM (58.0%) in the low-risk group had less than 3 partners in the past 12 months, while 59.0% of MSM who identified as being in the higher risk group had more than 10 partners in the same timeframe. When prompted about condom use, the rate of MSM in the low-risk group who reported “never use” when they have anal sex with male partners was higher than MSM (*p*< 0.05) in the high-risk group. Furthermore, regarding condom use with a casual male partner(s), the proportion of those choosing “yes” among the high-risk perception group was higher than people in the low-risk perception group (54.2% vs. 45.8%, *p*< 0.05). A higher number of MSM in the lower risk group selected “No” (57.3% vs. 42.7%, *p*< 0.05). The rate of substance use (ever using more than three substances) was higher in the high-risk group than the low-risk group and the difference was statistically significant (61.1% vs. 38.9%, *p*< 0.05). For participants who had ever been tested for an STI without any diagnosis, there was a higher proportion concentrated in the lower risk group than the higher risk group (56.8% vs. 43.2%, *p*< 0.05). People who have been diagnosed with an STI were concentrated more in the high-risk subgroup (66.7% vs. 33.3%, *p*< 0.05). HIV testing history was significantly different between the two groups. MSM in the high-risk group were more likely to have been tested for HIV in their lifetime and in the past year (*p*< 0.05).

### Factors of Self-Reported Risk Behaviors for High HIV Risk Perception

In bivariate analyses (**Error! Reference source not found**.), the following self-reported factors were associated with high risk perception compared with the low risk group: having more than 10 sexual partners in the past 12 months (OR=1.98, 95%CI: 1.19-3.32, *p*= 0.009), having had condom-less anal sex in the past 12 months (OR=1.59, 95%CI: 1.08-2.33, *p*= 0.017), and having used more than 3 substances (OR=1.56, 95%CI: 1.05-2.34, *p*= 0.029; OR=2.33, 95%CI: 1.18-4.87, *p*= 0.024).

### Barriers and Facilitators for HIV Testing

Bivariate analyses of HIV testing factors are shown in **Error! Reference source not found**.. The following are barriers: having married a female (OR=0.54, 95%CI:0.32-0.91, *p*= 0.019) and never or sometimes using condoms when having anal sex with male (OR=0.35, 95%CI:0.18-0.68, *p*= 0.002; OR=0.53, 95%CI:0.31-0.93, *p*= 0.025). Some facilitators were: higher risk perceptions (OR=2.60, 95%CI:1.68-4.01, *p*< 0.001), having 4 to 10 and more than 10 sexual partners in the past 12 months (OR=2.27, 95%CI:1.32-3.89, *p*= 0.003; OR=2.15, 95%CI:1.18-3.93, *p*= 0.013), knowledge of the last anal sex partner’s HIV status (OR=5.51, 95%CI:3.23-9.40, *p*= 0.025), and having been tested for STIs with and without a diagnosis (tested but without STI disease: OR=3.34, 95%CI:1.70-6.56, *p*< 0.001; diagnosed with one type of STI: OR=3.56, 95%CI:1.58-8.05, *p*= 0.002).

The factors significant in the bivariate analyses were included in multivariate analyses (**Error! Reference source not found**.). After adjusting for demographic characteristics, MSM who perceived themselves to be at a higher risk (AOR=3.42, 95%CI:2.06-5.68, *p*= 0.035), had 4 to 10 and more than 10 sexual partners in the past 12 months(AOR=2.60, 95%CI:1.40-4.83, *p*= 0.002; AOR=2.20, 95%CI:1.10-4.39, *p*= 0.025), knew their last anal sex partner’s HIV status (AOR=7.96, 95%CI:4.33-14.65, *p*<0.001), and have been tested for STI with and without a diagnosis(tested but without STI diagnosis: AOR=2.76, 95%CI:1.10-6.95, *p*=0.003; diagnosed with one type of STI: AOR=2.41, 95%CI:1.11-5.25, *p*<0.001), were more likely to have an HIV test in the past year. While never using and sometimes using a condom when having anal sex with a male were barriers for MSM to conduct HIV testing in the past 12 months (AOR=0.45, 95%CI:0.24-0.87, *p*=0.018; AOR=0.31, 95%CI:0.13-0.70, *p*=0.005).

## Discussion

In this study less than half of MSM identified themselves as having a high risk for HIV infection. Our findings illustrated that MSM with higher risk perceptions who have multiple anal sexual partners and knew their partner’s HIV status are more likely to participate in HIV testing. Seldom condom usage was a barrier for getting tested. Our study shows the importance of MSM to accurately perceive their own HIV infection risks, and also suggests interventions that contributed to reduce MSM’s sexual risk behaviors may indirectly increase HIV risk perception and promote HIV testing uptake.

Studies have found mixed levels of HIV risk self-perceptions among MSM. Previous surveys conducted in China[31] and the USA[32] reported that just a very small proportion of MSM have high-risk self-perceptions, even they had substantial-high risk[33]. In contrast, 57.54% of participants in Thailand[33] identified their own risk as high-level. Our study and previous research[34] have shown that MSM who engage in risky behaviors were more likely to evaluate themselves as belonging to the higher risk group. The number of different sexual partners (a behavior that is associated with risk) contributes greatly to accurate risk perceptions and HIV testing decisions in this study and has been explored in previous studies[35, 36]. The above findings also illustrate the close relationship between higher risk perceptions and HIV testing behavior. The result of condom use during anal sex is interesting in our study. MSM who had condomless anal intercourse with casual partners is significant in determining high-risk self-perceptions, while seldom condom use in general sex did not significantly contribute to higher risk perception for MSM but was rather associated with a lower likelihood for HIV testing. Data from Brazil showed a significant increase in the odds of perceiving a higher risk of HIV among MSM who had condomless anal intercourse in the past year[37]. Results from the USA illustrate that participants who perceived their risk of HIV as being low reported having condomless anal intercourse recently and were less likely to obtain HIV testing[38]. Previous studies have found mixed associations regarding condom use during sex, perceived risk, and HIV testing, possibly resulting from complex social and environmental factors associated with sexual behaviors and changes in sexual risk[39]. The inconsistency between risky sexual behaviors and self-perception of HIV risk may expose some degree of cognitive discordance [40] and lack of risk perception education[41] among MSM. Furthermore, the correlation between condom use and HIV testing also shows the importance of encouraging MSM who have additional risk factors (i.e. unprotected sex) to get tested for HIV regularly[42]. It also shows the importance of education regarding condom use[39]. Consistent with previous studies, those who know their partner’s HIV status and STI testing history (including STI diagnoses)[43, 44] were major facilitators for promoting HIV testing. Knowing a sexual partner’s HIV status or partner notification of HIV status before sex can be an important and feasible approach to increase HIV testing likelihood[45] and therefore reducing HIV infection among MSM[46]. China has already initiated partner notification programs[46] and tried to integrate STI testing with HIV testing in the clinic. In line with our study findings, this could be a valid way to help people identify their risk of HIV infection and to promote HIV testing[45].

Forming accurate risk perceptions among MSM and improving the frequency of HIV testing is important, but education may not be enough to reduce the risks of HIV[47]. MSM’s risk perceptions are influenced by multiple factors[48]. The IMB model recognizes that well-informed individuals are not motivated to change certain behaviors that can increase their HIV risk[49]. Culturally-informed and theory-driven risk education strategies are needed urgently[37]. The theory of triadic influence (TTI) is one of the most comprehensive and integrative theories of health behavior to date and is utilized by large numbers of people who are developing health promotion programs[50]. The TTI has been referenced as a helping guide in developing multiple health promotion programs for reducing substance use[51] to programs aimed at reducing risky behavior among adolescents[52]. TTI suggests that behavior change is correlated with various factors, including individual, social and environmental aspects. For future practical implications, nurses, trained peers, and other health prevention providers who have opportunities directly accessed to MSM could utilize this integrated theory to educate MSM about the risk associated with certain behaviors and encourage them to reduce high-risk behavior in the future[53]. After receiving risk reduction counseling, there may be a change in one’s risk perception and MSM may also begin to believe that they can make behavioral changes to gain better control over their HIV risk[47]. Moreover, intervention could focus on enhancing MSM’s rate of knowledge regarding sexual partners’ HIV status and therefore increase HIV testing proportions among MSM. Partner notification program (referring to the idea that sexual partners should take the initiative to tell each other their HIV infection status and encourage each other to get tested for HIV) as a public health measure was recommended by WHO[45]. This strategy with practical evidence[54, 55] has shown that notification services can increase HIV testing among partners of HIV-positive people[56, 57], and it could be integrated into routine counseling and HIV care follow up programs which led by nurses and other healthcare providers in CBO and HIV testing clinics. Ultimately, further research is needed to better understand the factors that shape HIV risk perception among MSM [58].

There are limitations in the present study. Only residents from Beijing were able to enroll in this study. This study does not include MSM who do not use BLUED or are outside Beijing, therefore it may not be generalizable to all MSM in China. Moreover, MSM’s risk perception was not specifically categorized and may not be accurately measured by just three items. Further population-specific and more reliable HIV risk assessment tools should be utilized. Also, the questionnaire’s categories of partner types can be unclear at times and may mislead people whose main partners are also casual. More accurate and well-defined options such as “regular” and “casual” partners should be provided in future research. Information on types of substance-using and the exact number of sexual partners were insufficient, and this restricts our ability to do further analysis on how different types of substance or multiple sex partners impact men’s risk perception. More details of sexual and substance use behavior should be collected in further study. Finally, self-reported data of sexual behaviors has inevitable bias, and it is unknown whether participants under-reported or over-reported their involvements in risky behaviors.

## Conclusion

In conclusion, over half of MSM in this study have high self-perceived HIV risk. We showed that self-perceived risk and other sexual behavioral factors were closely associated with HIV testing and emphasized the importance of enhancing HIV risk awareness and accessing HIV prevention practices among Chinese MSM. In today’s increasingly technological society, individualized intervention implemented through social media, or the internet would be more practical and problem-specific. Nurses, trained peers, and other healthcare providers are expected to lead design strategies with the support of technical sources, which eventually contribute to changing the sexual practices among those who perceive themselves to be at high risk of being infected with HIV. The developed intervention could focus on increasing condom use and awareness of each partners’ HIV infection status, and the social media-based intervention also allows for MSM individuals to avoid social and cultural stigma or even criminalization[59].

## Data Availability

The data underlying the results presented in the study are available requested from authors

